# Calibrating an SIR model for South Korea COVID-19 infections and predicting vaccination impact

**DOI:** 10.1101/2021.09.27.21264172

**Authors:** Brandon Pae

## Abstract

In the span of 1.5 years, COVID-19 has caused more than 4 million deaths worldwide. To prevent such a catastrophe from reoccurring, it is necessary to test and refine current epidemiological models that impact policy decisions. Thus, we developed a deterministic SIR model to examine the long-term transmission dynamics of COVID-19 in South Korea. Using this model, we analyzed how vaccines would affect the number of cases. We found that a 70% vaccination coverage with a 100% effective vaccine would effectively eliminate the number of cases and herd immunity would have been obtained approximately 85 days after February 15 had there not been a reintroduction of cases.

## Introduction

With over 190 million global COVID-19 cases, international governments and health organizations have worked to implement health policies, including quarantine measures, to effectively reduce COVID-19 transmissions and subsequent deaths. Nonuniform COVID-19 response has produced varying results in different countries. While inadequate testing and quarantine measures led to surging cases in countries like the United States, in countries like South Korea the government quickly distributed testing, employed extensive contact tracing, and provided efficient treatments, leading to a rapid reduction in the number of COVID-19 cases.

South Korea’s first case of COVID-19 was reported on January 20, 2020. After the number of cases started to rise in February, the government quickly shipped thousands of kits each day to institutions and developed many screening centers, which helped to rapidly reduce the number of infections. Treatment centers were also established to both quarantine and treat COVID-19 patients. Furthermore, the government improved contact tracing by increasing the number of contact tracers, providing them with more data, and promoting citizen assistance. These efficient and rapidly implemented policies severely reduced infections and made South Korea a great model for other countries.

## Methods

To model South Korea’s early COVID-19 situation, we initially developed an SEIR model instead of an SIR model. Our SEIR model had five compartments: Susceptibles (S), Exposed (E), Infected and Symptomatic (Is), Infected and Asymptomatic (Ia), and Recovered (R). This model had the following parameters: β (the infection rate), γ (the recovery rate), λ (the force of infection), k (the reciprocal of the latent period), p (the percentage of infections that are symptomatic), b (the birth rate), and µ (the background mortality rate).

However, to solve the differential equations at each timestep, we used RStudio’s ode function, which is optimal for only three compartments instead of five. As a result, we had to make some assumptions to simplify the SEIR model. First, we assumed that symptomatic and asymptomatic infections had the same recovery rate, meaning that we could combine Is and Ia into one compartment, I. Second, we assumed that there was no latent phase, so any susceptibles who were exposed immediately became infectious. From these assumptions, we simplified the SEIR model into an adapted SIR model.

From Worldometer, we obtained the South Korea data for daily active COVID-19 cases from February 15, 2020, to December 4, 2020. We confirmed the validity of this data by first comparing it to the data from the AMCHAM Border Member Companies, for which we subtracted the total number of removed cases (deaths and recoveries) from the total number of infected cases and obtained the same values for the number of active infected cases on a given day. This Worldometer data can be further validated using the World Health Organization’s data. Since WHO reports new South Korea cases by week, it is necessary to calculate 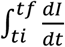 dt for each week, where tf is the final day and ti is the first day. This integral is the number of new infected cases in each week.

In our adapted SIR model, the independent variable is the time elapsed since February 15 in units of days. The dependent variable is the prevalence of infected people, symptomatic and asymptomatic, in South Korea. With the initial number of susceptible, infected, and recovered people on February 15, we utilized ode() to solve the differential equations and find the number of susceptible, infected, and recovered individuals on each subsequent day. However, before fitting an SIR model to the South Korea case data, we needed to find appropriate values for the two unknown parameters: beta (the infection rate) and gamma (the recovery rate). We employed the dpois method to calculate how well an SIR model (with certain values of beta and gamma) matched the South Korea case data. In contrast to the least sum of squares approach, this likelihood method includes a range of uncertainty for the parameter values and enables the integration of the reporting rate into the calculation. Then, using the optim() calibration method, we found the values for beta and gamma that generated the best likelihood, or fit, for the Worldometer data.

We separated the infected case data into three portions, and calibrated models with unique parameter values for each section. The reason for this choice is because we cannot develop a single SIR model with constant parameters for the entirety of the 294 days. Such a model will only rise and decline once, because as more susceptibles become infected, it becomes harder for the pathogen to spread, and the epidemic eventually declines. However, because many new people entered South Korea multiple times, there were multiple surges in infected cases. As a result, our initial SIR model, as pictured in Figure 2, became very inaccurate. Thus, we created three individual models with separate parameters for the data, as shown in Figure 3.

**Figure 1:**
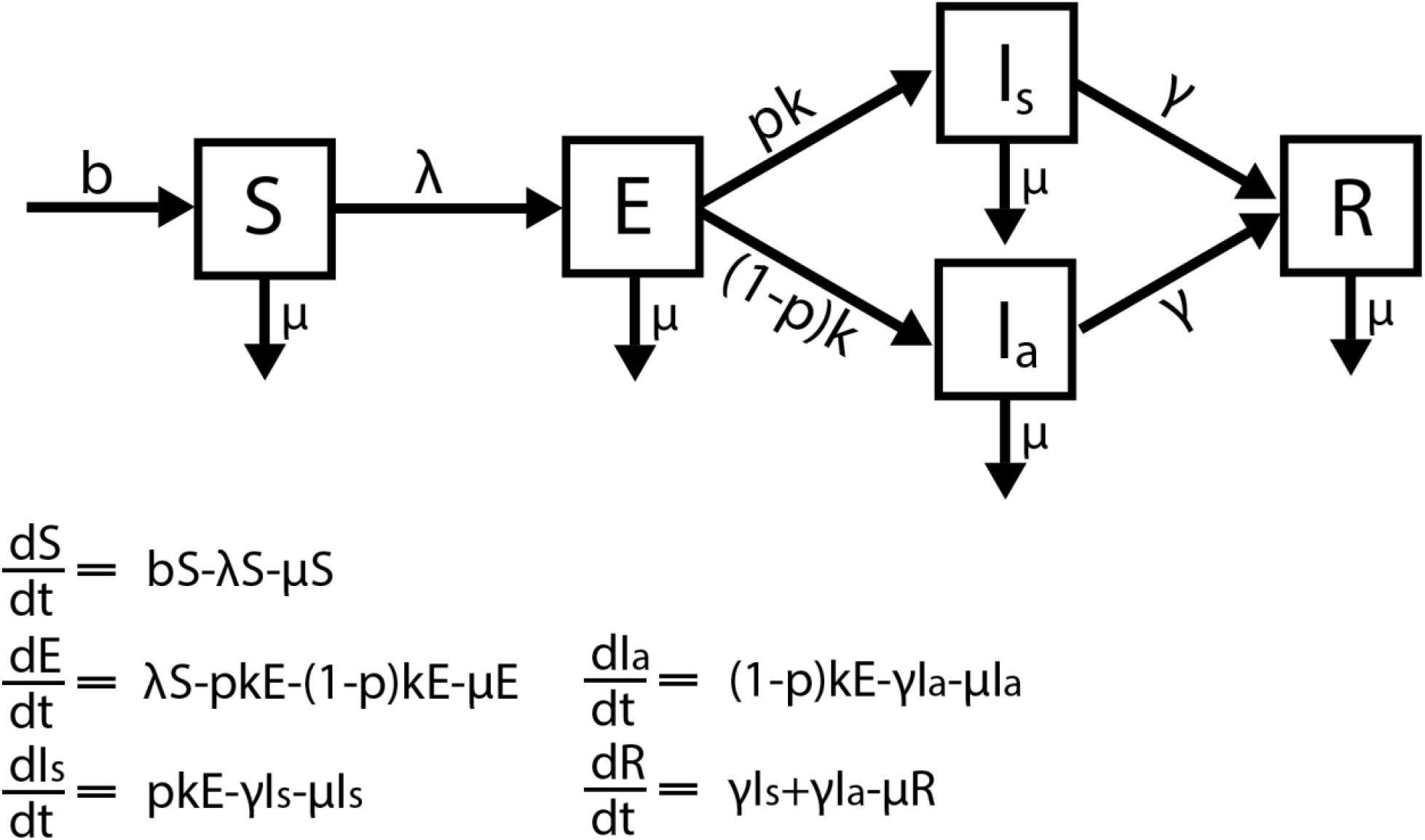
A diagram of our SEIR model and its differential equations.

**Figure 2:**
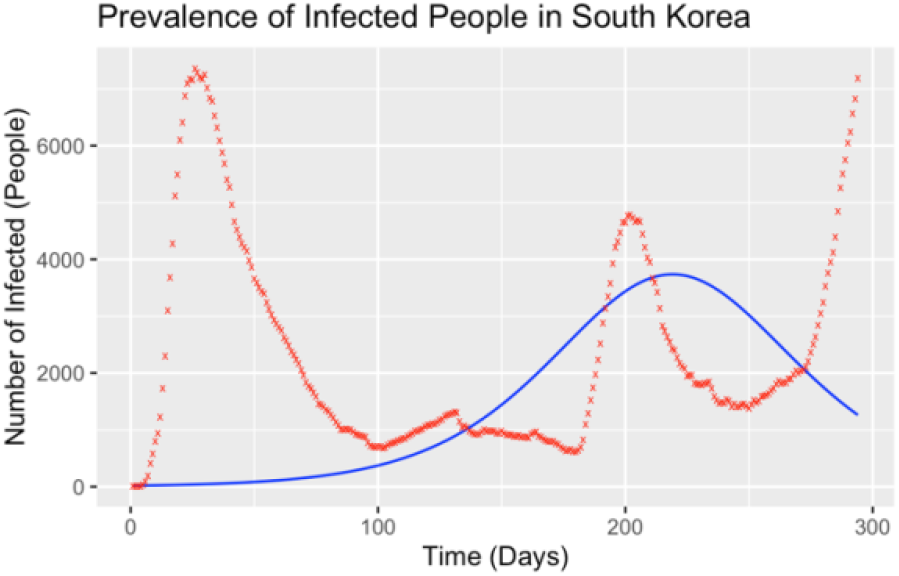
Calibrated Model using Likelihood.

**Figure 3:**
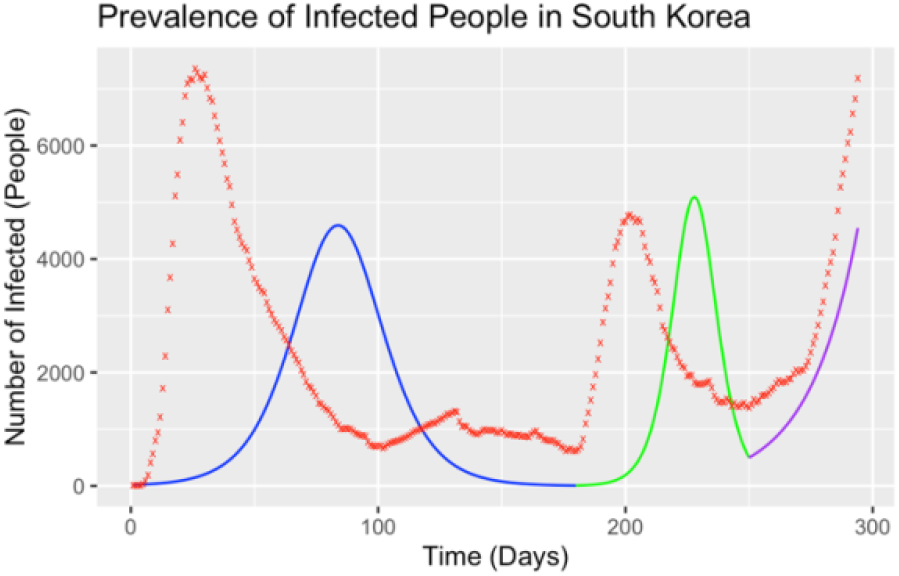
Updated Calibrated Model.

After implementing our SIR model, we predicted the effects of a vaccine. Since vaccines from Pfizer and Moderna are 95% effective, we estimated how a vaccine with 100% efficacy would impact the number of infections. We introduced the parameter p, which represents the vaccination coverage or the percentage of the population that has been vaccinated. Then, we simulated instantaneous vaccination of p percent of the population 250 days after February 15, which is when the last surge in cases began in Figure 3.

Our full code can be found here: https://github.com/paeb/Covid19-Research

## Results

In the SIR model shown in Figure 3, the average value for beta was 6.25, and the average value for gamma was 6.04. The recovery rate was only slightly less than the infection rate, which explains why the rise in cases was more gradual in South Korea than in other countries. Our SIR model predicts that herd immunity would have been obtained around 85 days after February 15, since the number of secondary infections caused by a single case, Reff, is equal to 1 at that time. However, because of the multiple influxes of infected people, this prediction was inaccurate.

Without any vaccination, the SIR model is as pictured in Figure 3. However, when 5% of the population has been vaccinated, the model is as shown in Figure 5. When 70% of the population has been vaccinated, the epidemic is estimated to fall within 20 days, as shown in Figure 4.

**Figure 4:**
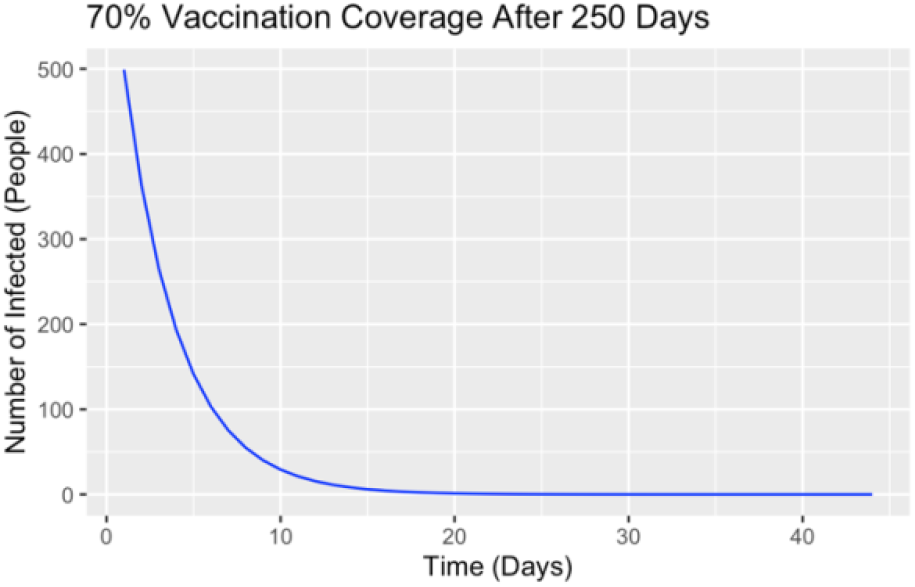
70% of the population is vaccinated.

**Figure 5:**
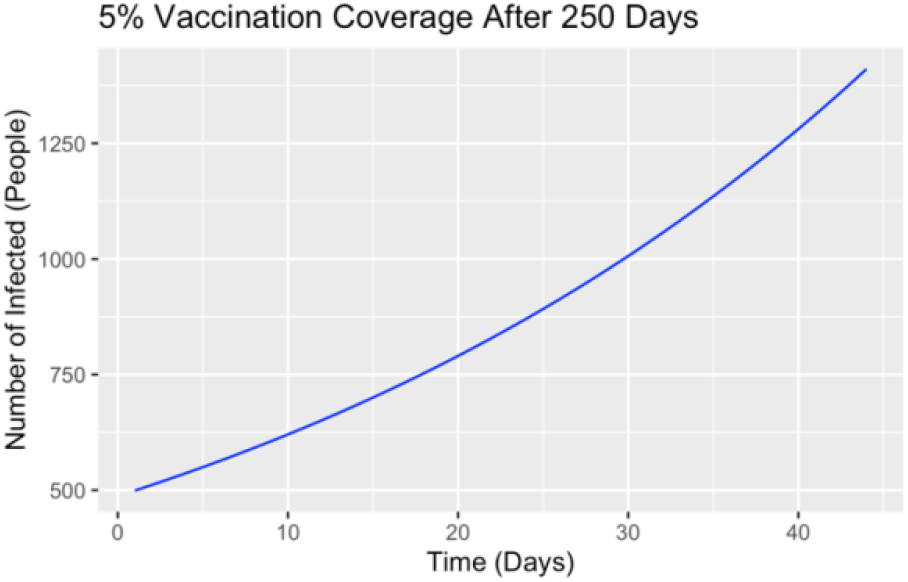
5% of the population is vaccinated.

## Discussion

Since the current SIR model parameters do not give a close enough fit to the observable data, as shown in Figure 3, we need to include more competing hazard rates to make a more accurate model. One major error in our model is homogeneity: we assume that the hazard rates affect each member of each compartment equally but based on population structure and unique interactions between different groups of people, this is not valid. With more specific case data, we could develop an age-stratified model and have separate hazard rates for each age group, which could augment the SIR model’s accuracy. Additionally, our model assumed that vaccination was instantaneous 250 days after February 15, but vaccination is a lengthy process that often lacks full compliance in treatment, so the effects were not as predicted.

By addressing these flaws, we can implement more accurate SIR models and better predict the course of an epidemic. These models can effectively determine which policy measures will be the most impactful and prevent the epidemic from claiming more lives.

## Data Availability

The data was acquired from the dataset on Worldometer

https://www.worldometers.info/coronavirus/country/south-korea/

